# Association between the Epigenetic Lifespan Predictor GrimAge and History of Suicide Attempt in Bipolar Disorder

**DOI:** 10.1101/2022.11.15.22282309

**Authors:** Camila N. de Carvalho Lima, Emese H.C. Kovács, Salahudeen Mirza, Alexandra Del Favero-Campbell, Alexandre Paim Diaz, Joao Quevedo, Benney M.R. Argue, Jenny Gringer Richards, Aislinn Williams, John A. Wemmie, Vincent A. Magnotta, Jess G. Fiedorowicz, Jair C. Soares, Marie E. Gaine, Gabriel R. Fries

**Affiliations:** Translational Psychiatry Program, Faillace Department of Psychiatry and Behavioral Sciences, The University of Texas Health Science Center at Houston. 1941 East Rd, 77054 Houston, TX; Department of Neuroscience and Pharmacology, The University of Iowa. 51 Newton Rd, 52242 Iowa City, IA; Institute of Child Development, University of Minnesota. 51 E River Rd, 55455 Minneapolis, MN; Center for the Study and Prevention of Suicide, Department of Psychiatry, University of Rochester Medical Center, Rochester, NY; Center of Excellence in Mood Disorders, Faillace Department of Psychiatry and Behavioral Sciences, The University of Texas Health Science Center at Houston. 1941 East Rd, 77054 Houston, TX; Neuroscience Graduate Program, The University of Texas MD Anderson Cancer Center UTHealth Graduate School of Biomedical Sciences, Houston, TX. 6767 Bertner Ave, 77030 Houston, TX; Pharmaceutical Sciences and Experimental Therapeutics, The University of Iowa. 180 South Grand Ave, 52242 Iowa City, IA; Department of Radiology, The University of Iowa. 200 Hawkins Dr, 52242 Iowa City, IA; Department of Psychiatry, The University of Iowa. 200 Hawkins Dr, 52242 Iowa City, IA; Iowa Neuroscience Institute, The University of Iowa. 169 Newton Rd, 52242 Iowa City, IA; University of Ottawa Brain and Mind Research Institute, Ottawa Hospital Research Institute. 501 Smyth, K1H 8L6, Ottawa, ON; Center for Precision Health, School of Biomedical Informatics, The University of Texas Health Science Center at Houston, Houston, TX, USA. 7000 Fannin, 77030 Houston, TX

**Author notes:** **Corresponding Author:** Gabriel R. Fries, PhD, 1941 East Rd, 77054 Houston, TX. Shared first-authorship (Camila N. de Carvalho Lima, Emese H.C. Kovács). Shared last authorship (Marie E. Gaine, Gabriel R. Fries).

## Abstract

Bipolar disorder (BD) has been previously associated with premature mortality and aging, including acceleration of epigenetic aging. Suicide attempts (SA) are greatly elevated in BD and are associated with decreased lifespan, biological aging, and poorer clinical outcomes. We investigated the relationship between GrimAge, an epigenetic clock trained on time-to-death and associated with mortality and lifespan, and SA in two independent cohorts of BD individuals (discovery cohort - controls (n=50), BD individuals with (n=77, BD/SA) and without (n=67, BD/non-SA) lifetime history of SA; replication cohort - BD/SA (n=48) and BD/non-SA (n=47)). An acceleration index for the GrimAge clock (GrimAgeAccel) was computed from blood DNA methylation (DNAm) and compared between groups with multiple general linear models. Differences in epigenetic aging from the discovery cohort were validated in the independent replication cohort. In the discovery cohort, controls, BD/non-SA, and BD/SA significantly differed on GrimAgeAccel (*F=*5.424, *p*=0.005), with the highest GrimAgeAccel in BD/SA (*p*=0.004, BD/SA vs. controls). Within the BD individuals, BD/non-SA and BD/SA differed on GrimAgeAccel in both cohorts (*p*=0.008) after covariate adjustment. Finally, DNAm-based surrogates revealed possible involvement of plasminogen activator inhibitor 1, leptin, and smoking pack-years in driving accelerated epigenetic aging. These findings pair with existing evidence that not only BD, but also SA, may be associated with an accelerated biological aging and provide putative biological mechanisms for morbidity and premature mortality in this population.

## 1. Introduction

Bipolar disorder (BD) is a severe and chronic psychiatric disorder with an estimated global prevalence of at least 1% [1-2]. Aside from significant functional impairments in daily life [3], BD is associated with premature mortality likely due to comorbid medical conditions including cardiovascular diseases, diabetes mellitus, obesity, endocrine and thyroid diseases [4], and suicide [5]. On average, the lifespan of individuals with BD is 9-17 years shorter than that of the general population [6,7]. The suicide rate for BD is 10-30 times higher than the general population, with 20-60% of these individuals attempting suicide at least once in their lifetime [8]. Additionally, suicide attempt (SA) is associated with decreased lifespan, which is accounted for by, besides an increased risk of suicide, comorbid medical conditions [9].

Emerging biomarkers of aging have begun to clarify the observed shortened lifespan in BD and, to a lesser extent, SA. DNA methylation (DNAm) varies across the lifespan, involves both genetic and environmental contributions, and can reversibly modulate gene expression [10]. DNAm varies with age, both globally and locally, and is therefore an appealing measure from which to derive estimates of aging more closely tied to biological development [11], contrary to the weaker predictor, time-since-birth [12]. Epigenetic clocks, which estimate age from DNAm patterns, offer biologically-derived age estimates which can be directly compared to chronological age to determine acceleration or deceleration of biological aging [13]. These clocks have traditionally been trained on chronological age data [14,15] and have been associated with various neurodegenerative and psychiatric diseases [16,17], including BD [18, 19]. As chronological age is not perfectly synonymous with lifespan, novel epigenetic clocks specifically trained on phenotypical age and lifespan data are beginning to emerge. GrimAge, a clock based on DNAm-based predictors of plasma proteins and a DNAm-based smoking score, was specifically trained on time-to-death and has outperformed others when predicting all-cause mortality [20]. GrimAgeAccel, a measure derived from the regression of DNAm-based age on chronological age, provides information on accelerated (positive) or decelerated (negative) epigenetic aging (EA). Of note, positive GrimAgeAccel has been associated with deficits in neurocognitive and neurostructural outcomes, such as lower cognitive ability and vascular brain lesions [21].

Although investigations of EA using first-generation clocks have revealed accelerated aging in BD [18,19], the mortality-associated GrimAgeAccel has only recently focused on BD, with work by our group identifying higher GrimAgeAccel in BD as well as its relationship with cognitive impairment in BD [22]. Given the evidence that suicidal behavior, aside from the heightened mortality associated with suicide, also associates with premature mortality for natural causes [9], EA may be a plausible biological candidate for this risk elevation. EA studies in suicidal behavior have documented acceleration across measures [23-25], but the only study that investigated GrimAgeAccel in the context of suicide, comparing groups of high and low lethality SA, found no significant difference [26].

No study has considered GrimAgeAccel in BD or a matched-diagnosis reference group with and without SA. This consideration is critical to not only advance the understanding of suicide within BD specifically, but to clarify whether EA associated with SA is specific to SA *vs*. BD diagnosis. Furthermore, studying SA within BD allows for rigorous insights to uniquely associated biological mechanisms, as reference groups in previous studies have often been non-psychiatric controls. In this study, we investigated whether GrimAgeAccel differentiates individuals with BD and a history of SA (BD/SA) from those with BD and no history of SA (BD/non-SA). We leverage a richly phenotyped discovery cohort with an independent replication cohort to test whether GrimAgeAccel is especially pronounced in BD/SA.

## 2. Materials and Methods

### 2.1 Discovery cohort

Sample recruitment for this study has been recently described [22]. Briefly, 144 BD (124 BD-I/20 BD-II) individuals were recruited alongside 50 non-psychiatric controls (CON) matched for age, sex, and race at the Center of Excellence in Mood Disorders, Houston, TX. BD diagnosis and features of illness severity (substance use comorbidity, total number of comorbidities, total number of psychiatric hospitalizations, age of onset for mood disorder, length of illness) were ascertained in the Structured Clinical Interview for DSM-IV Axis I Disorders (SCID-I), and a standardized method was used to collect demographic information. Young Mania Rating Scale (YMRS) [27] and Montgomery-Åsberg Depression Rating Scale (MADRS) [28] were administered. The criterion for suicidality was one or more documented actual, aborted, and interrupted SA assessed by the Columbia Suicide History Form (CSHF) [29] yielding subgroups of BD/SA (n=67) and BD/non-SA (n=77). Exclusion criteria included other medical conditions such as neurological disorders, traumatic brain injury, current pregnancy, schizophrenia, developmental disorders, eating disorders, and intellectual disability. CON (n=50) had neither history of any Axis I disorder in first-degree relatives nor prescribed psychotropic medication. Clinical interviews were administered by trained evaluators and reviewed by a board-certified psychiatrist. All participants completed a urine drug screen to exclude current illegal drug use. Informed consent was obtained from all participants at enrollment and prior to any procedure, and the protocol for the study was approved by the local institutional review board (IRB, HSC-MS-09-0340).

### 2.2 Replication cohort

Subjects were recruited through the Iowa Neuroscience Institute Bipolar Disorder Research Program of Excellence (BD-RPOE), approved by the IRB of the University of Iowa (IRB#201708703) [30-36]. Participants between 18 and 70 years with the ability to consent and a confirmed DSM-IV diagnosis of BD-I were recruited. History of actual SA and number of lifetime attempts were recorded with the Columbia Suicide Severity Rating Scale [37], a rating scale which definitions of suicide behavior were based in the CSHF used in the discovery cohort. Illness severity features were ascertained with a standardized method to collect demographic information and the Mini-International Neuropsychiatric Interview [38]. YMRS [27] and MADRS [28] were administered. Exclusion criteria included a history of loss of consciousness for more than 10 minutes, seizure disorder, brain damage or other neurological problems, coronary or cerebral artery disease, alcohol or drug dependence within the past 3 months, current pregnancy, or contraindication for magnetic resonance imaging were also excluded. The final replication cohort included 48 BD/SA and 47 BD/non-SA and did not include CON.

### 2.3 DNA extraction

Blood was collected by venipuncture in EDTA-containing vacutainers and stored at −80°C. In the discovery cohort, buffy coat from fasting participants was isolated before storage, followed by DNA isolation with the DNeasy Blood & Tissue Mini Kit (Qiagen, Hilden, Germany) and quantification on NanoDrop (Thermo, Waltham, MA, USA). In the replication cohort, 1 mL of whole blood per sample from non-fasting participants was used with the Puregene Blood Kit with RNase A solution (Qiagen). Elution Buffer CDB-02 (Kurabo Industries Ltd, Osaka, Japan) was used instead of DNA Hydration Solution.

### 2.4 Methylation assay

Five hundred nanograms of DNA were bisulfite-converted with the EZ DNA Methylation Kit (Zymo Research, Irvine, CA, USA) and interrogated for genome-wide DNAm on the Infinium EPICMethylation BeadChip (Illumina, San Diego, CA, USA). Poor quality probes with detection *p*-values <0.01 were excluded using *minfi* [39]. From the DNAm data, estimates of white blood cell count proportions (CD8+ T-lymphocytes (CD8T), CD4+ T-lymphocytes (CD4T), monocytes (Mono), granulocytes (Gran), natural killer cells (NK), and B-lymphocytes (B cell)) using the Houseman procedure [40] and smoking scores using EpiSmokEr [41,42] were retrieved. DNAm-based calculations of GrimAge, plasminogen activation inhibitor 1 (PAI-1), growth differentiation factor 15 (GDF-15), leptin, tissue inhibitor metalloproteinases 1 (TIMP1), cystatin C, adrenomedullin (ADM), beta-2-microglobulin (B2M), and smoking pack-years (PACKYRS) were performed using the New DNA Methylation Age Calculator (https://dnamage.genetics.ucla.edu/). GrimAgeAccel and age-adjusted values for the other DNAm clocks were calculated by regressing the predicted epigenetic ages on chronological age and using the residuals as acceleration indices. Positive and negative GrimAgeAccel values represent acceleration and deceleration of GrimAge, respectively.

### 2.5 Genotyping

Samples from the discovery cohort were genotyped on the Infinium Global Screening Array-24 (Illumina). Principal components (PC) analysis was performed, and the first three PCs were retained for use as covariates of genomic ancestry.

### 2.6 Statistical analyses

All variables included as outcomes in analyses were examined for normality with the Shapiro– Wilk test and, if necessary, log-normalized. For covariate analyses, genomic data were not available for the replication cohort. Instead, self-reported race was used as a substitute for PCs.

In both cohorts, the association between GrimAge and chronological age was assessed within each group using Spearman’s correlation test **(Figures 1A-B and S1)**. For demographic **(Table 1)** and clinical **(Table S1)** variables, Wilcoxon rank sum test with continuity correction, independent samples *t*-test, one-way analysis of variance (ANOVA), and Kruskal-Wallis tests were applied to reveal between-group differences. A comparison of the two cohorts was conducted with Fisher’s Exact Test and Welch Two Sample *t*-test/Wilcoxon **(Table S2)**. Analysis of covariance (ANCOVA) models were tested in the discovery cohort to predict GrimAgeAccel from group (**Table S3 and 2**). In Model 1, covariates included age, sex, years of education, and PCs. In Model 2, covariates were age, sex, years of education, PCs, and body mass index (BMI). In Model 3, covariates were age, sex, years of education, PCs, BMI, and white blood cell count proportions. In Model 4, covariates were age, sex, years of education, PCs, BMI, white blood cell count proportions, and smoking score. Finally, in Model 5, smoking score was the only covariate. This set of analyses was not repeated in the replication cohort due to the lack of a control group. *p*-values < 0.01 were considered statistically significant after Bonferroni correction for multiple testing.

**Table 1.**
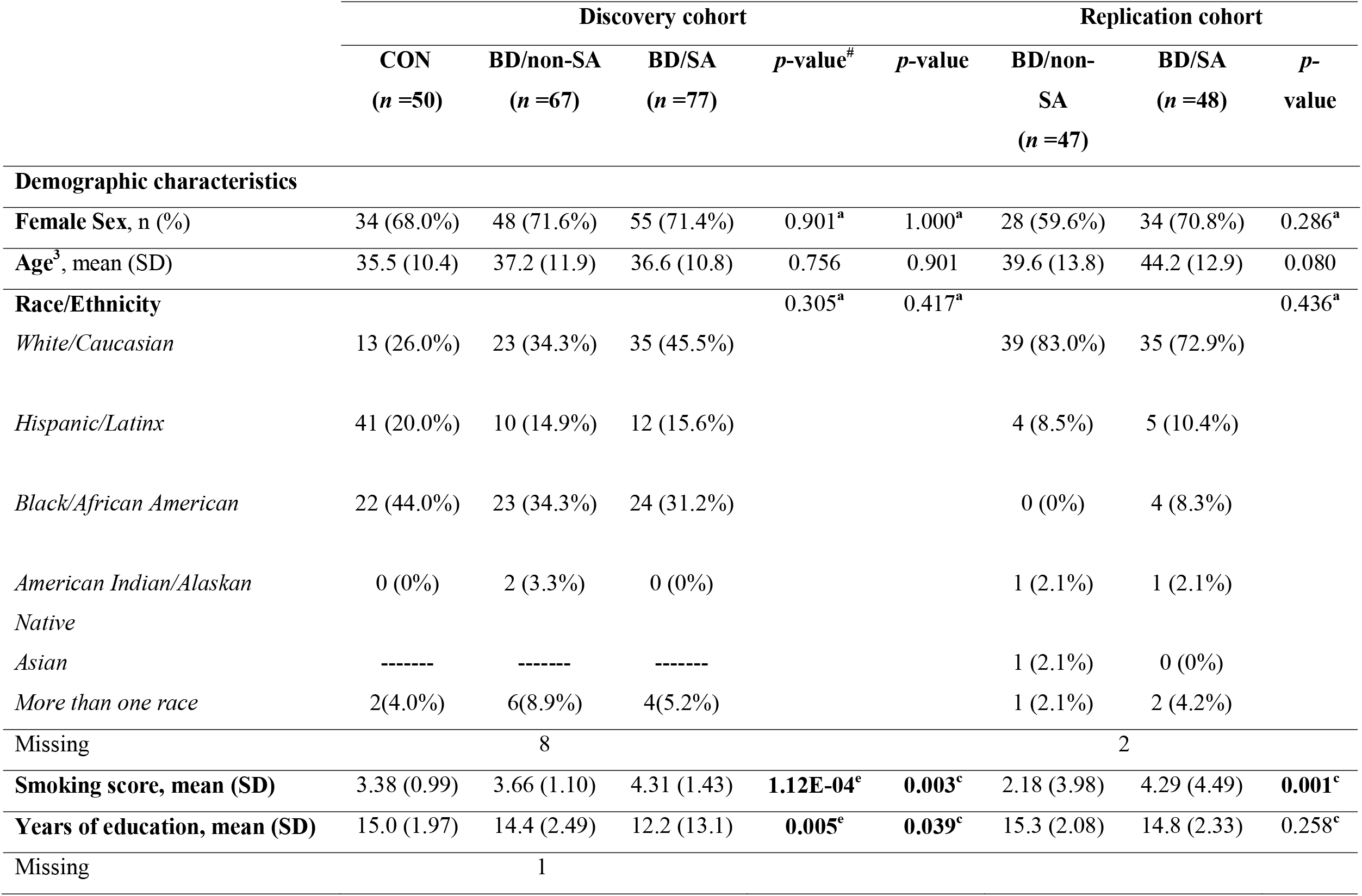
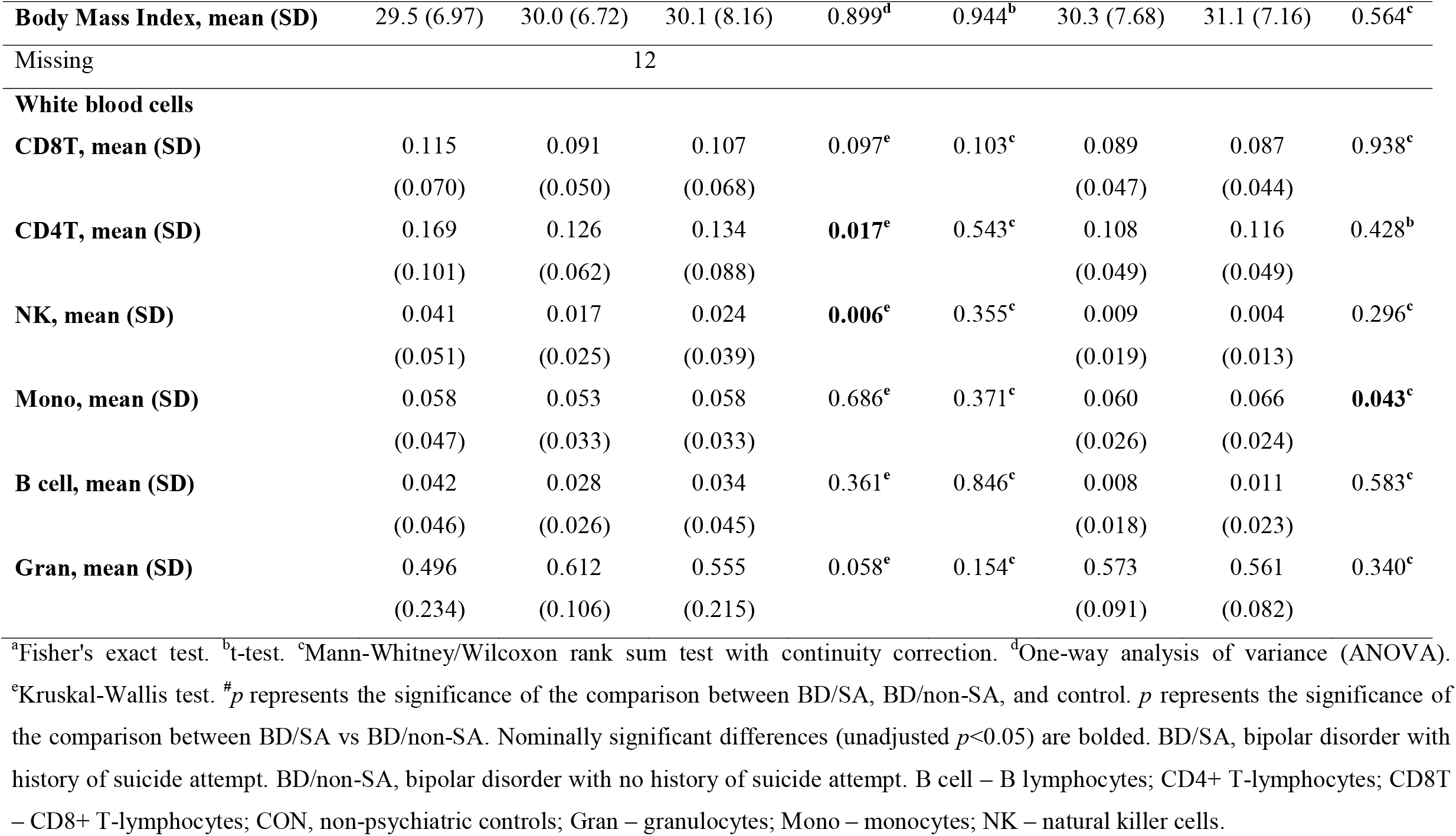
Demographic information.

**Figure 1.**
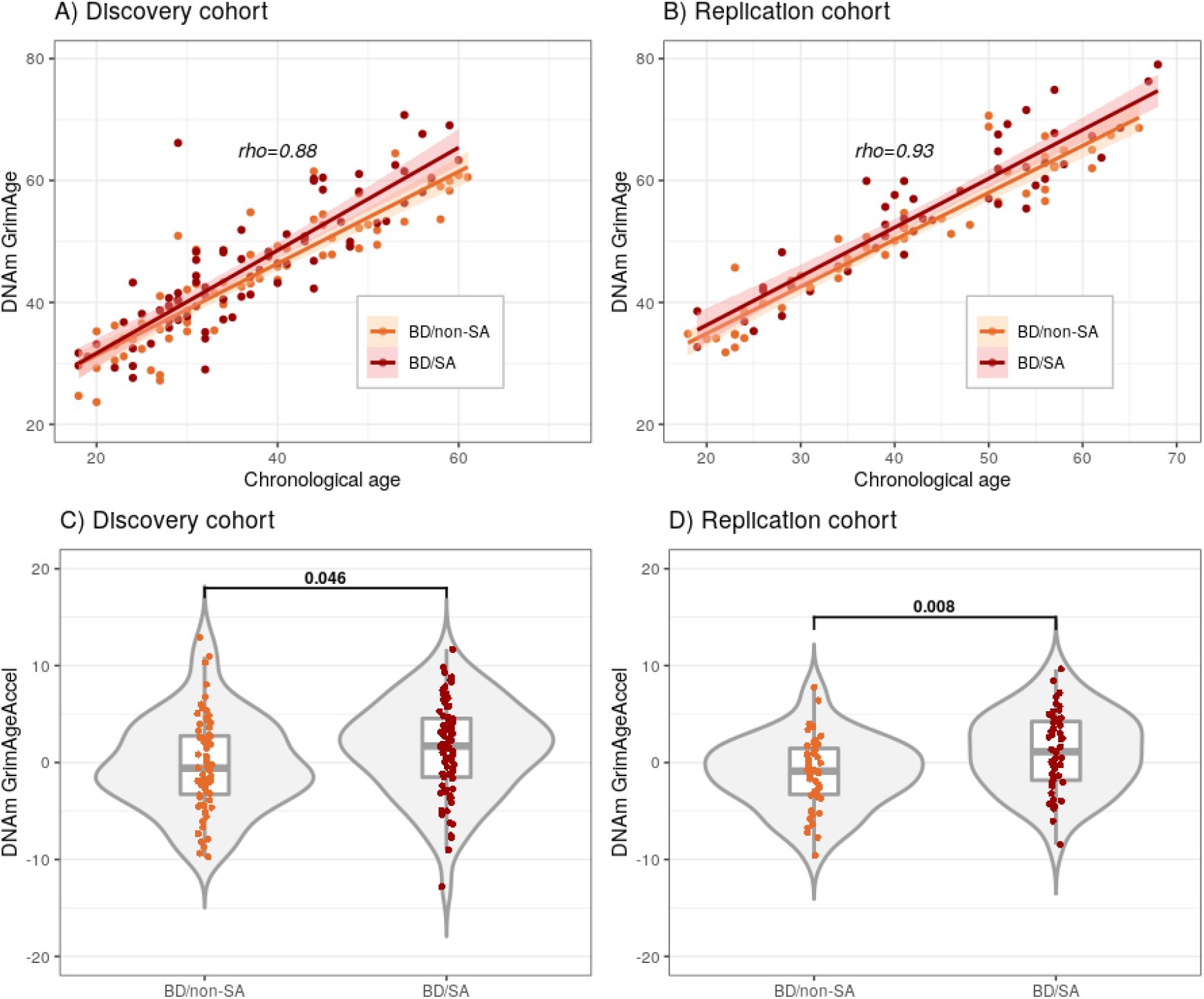
Accelerated DNAm GrimAge and GrimAge in BD groups. **A and B)** Correlation between estimated GrimAge and chronological age in **A)** the discovery cohort. Spearman’s correlation analysis indicates a significant correlation (*p<*2.2E-16) between DNAm GrimAge and chronological age in the entire sample, as well as in the subgroups (BD/non-SA: rho=0.91, *p*<2.2E-16; BD/SA: rho=0.85, *p*<2.2E-16). **B)** In the replication cohort, Spearman’s correlation analysis indicates a significant correlation (*p*<2.2E-16) between DNAm GrimAge and chronological age in the entire sample, as well as in the subgroups (BD/non-SA: rho=0.95, *p*<2.2E-16; BD/SA: rho=0.90, *p*<2.2E-16). **C and D)** DNA methylation (DNAm) accelerated GrimAge (GrimAgeAccel, log-normalized) in BD/non-SA and BD/SA in **C)** the discovery cohort (n=67 BD/non-SA and n=77 BD/SA) and **D)** the replication cohort (n=47 BD/non-SA and n=48 BD/SA). *p*-values determined by independent samples t-tests. BD/SA, bipolar disorder with history of suicide attempt. BD/non-SA, bipolar disorder with no history of suicide attempt.

In the discovery cohort, two models were tested to predict each DNAm-based, normalized, epigenetic clock per group **(Table S4)**. Model 6 was an ANOVA and in Model 7 (ANCOVA), age, sex, PCs, and years of education were included as covariates.

In both the discovery and the replication cohort, these models were tested to predict each DNAm-based, age-adjusted epigenetic clock per group (BD/non-SA, BD/SA) **(Table 3)**. Model 6 was repeated with Welch’s independent samples *t*-test to compare the two groups. Finally, to investigate the potential mechanisms for elevated GrimAgeAccel in BD/SA compared to BD/non-SA, we tested six dichotomic logistic regression models to predict GrimAgeAccel in each cohort, including severity variables as covariates **(Table 4)**. As outcome in the analyses, data were divided by percentile of GrimAgeAccel into two groups (high and low GrimAgeAccel). Covariates included length of illness, total number of psychiatric comorbidities, substance use comorbidity, age of onset for mood disorder, current lithium use (yes/no), and prescribed medication use (yes/no). All analyses were performed in R 4.1.1 [43], using *ggplot2* [44], *tidyr* [45], *dplyr* [46], *magrittr* [47], and *ggpubr* [48] packages, as well as SPSS 28[49].

**Table 2.** Adjusted analysis of differences in GrimAgeAccel between BD/non-SA and BD/SA groups.

|  | Discovery cohort |  | Replication cohort<sup>a</sup> |  |
| --- | --- | --- | --- | --- |
| | $F_{\text{BD/SA}}$ (df, df=1,) | $p$-value | $F_{\text{BD/SA}}$ (df, df=1,) | $p$-value |
| **Model 1** | 5.126 | **0.025** | 7.683 | **0.007** |
| **Model 2** | 3.896 | 0.051 | 8.654 | **0.004** |
| **Model 3** | 4.137 | **0.044** | 8.837 | **0.004** |
| **Model 4** | 5.527 | **0.020** | 17.000 | **9.55E-05** |
| **Model 5** | 5.127 | **0.025** | 10.040 | **0.002** |

Model 1: covaried for age, sex, GWAS principal components (PCs), and years of education. Model 2: covaried for age, sex, GWAS, PCs, years of education, and body mass index (BMI). Model 3: covaried for age, sex, GWAS PCs, years of education, BMI, and white blood cell proportions. Model 4: covaried for age, sex, GWAS PCs, years of education, BMI, white blood cell proportions, and smoking score. Model 5: covaried for smoking score. Bonferroni-corrected $p < 0.01$ was taken as the significance threshold. Nominally significant differences ( $p < 0.05$ ) are bolded. <sup>a</sup>Used race instead of genomic ancestral PCs. BD/SA, bipolar disorder with history of suicide attempt. BD/non-SA, bipolar disorder with no history of suicide attempt.

**Table 3.**
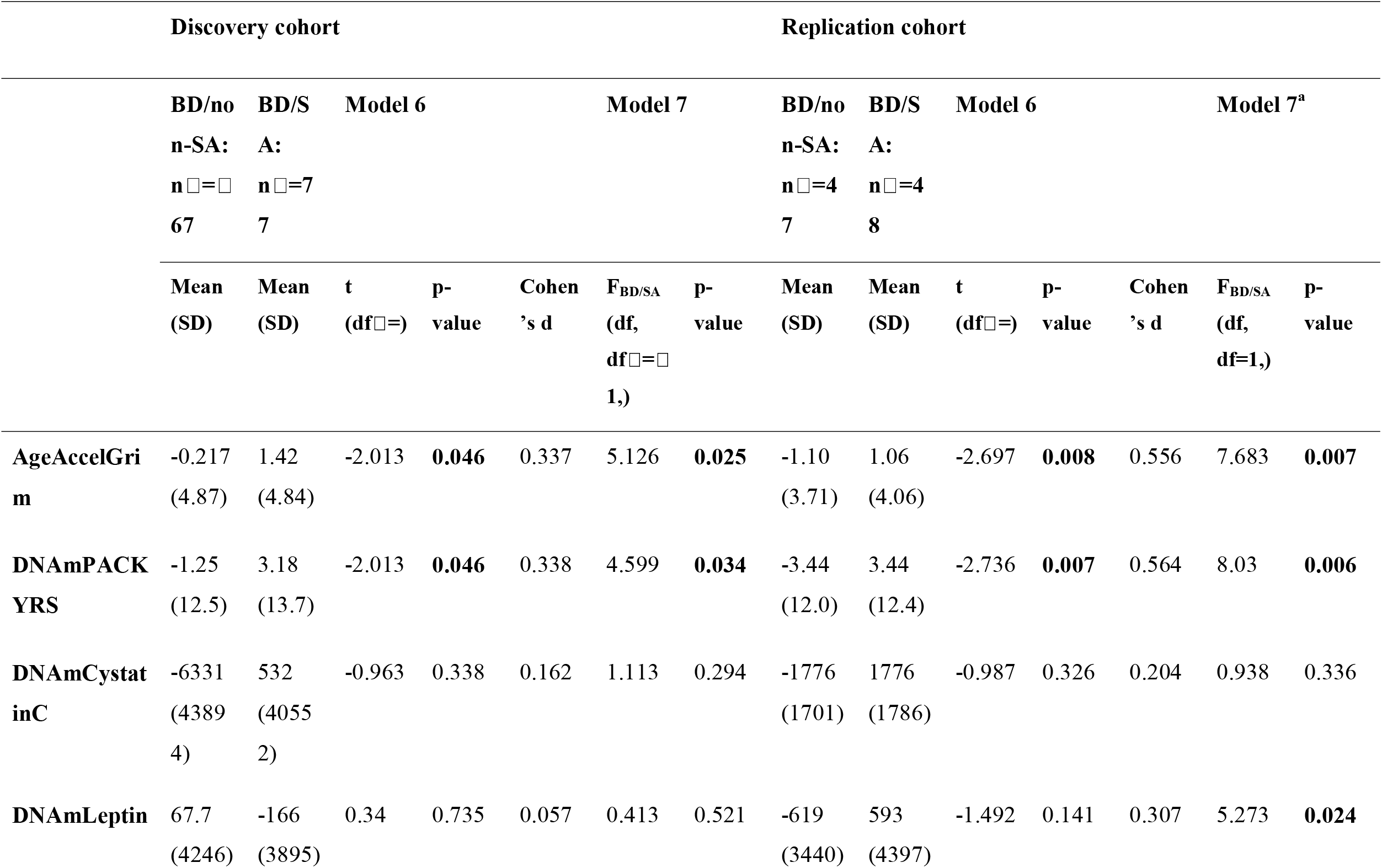
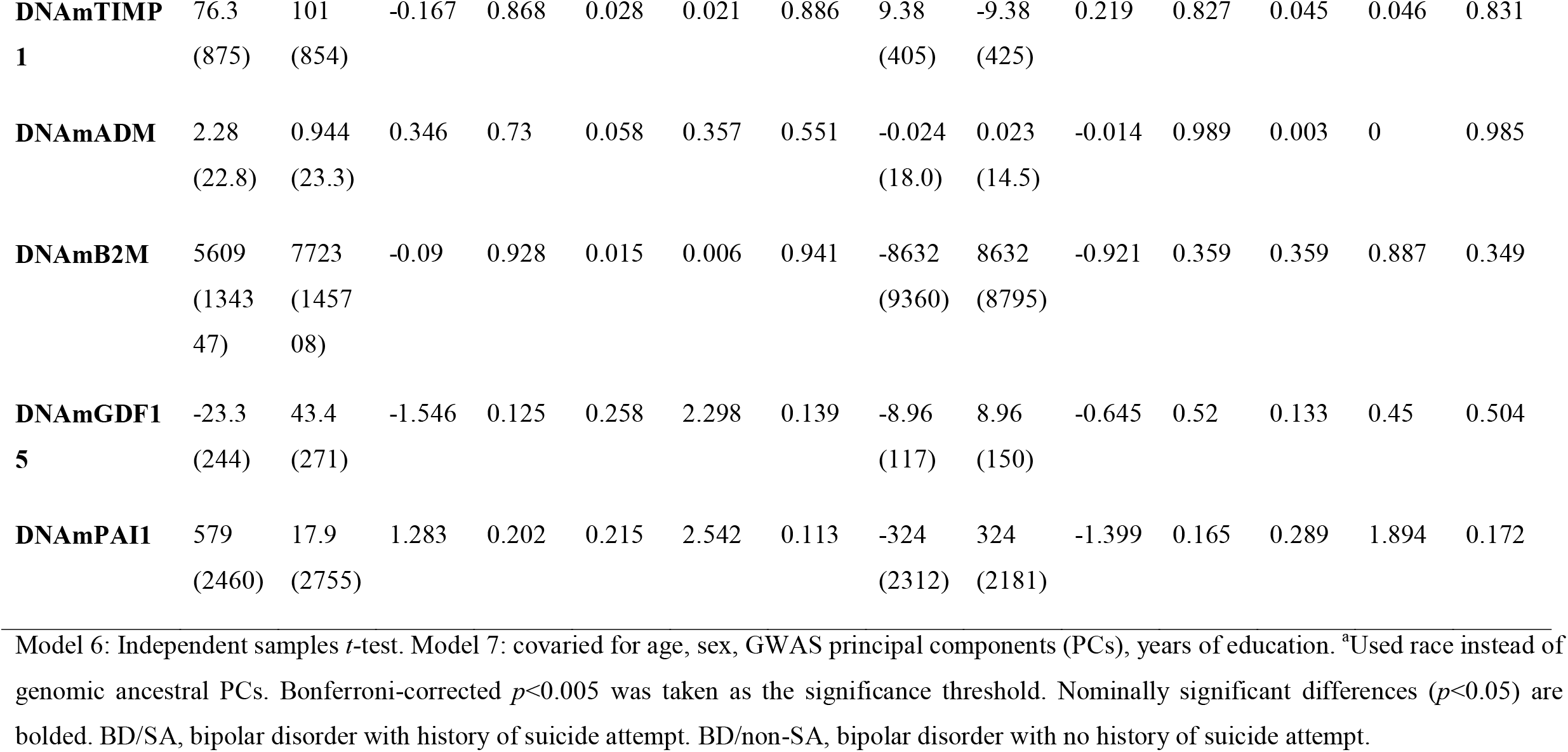
GrimAge Clock and its components in BD/non-SA and BD/SA.

**Table 4.** Phenotype-adjusted differences in GrimAgeAccel between BD/non-SA and BD/SA groups.

|  | Discovery cohort |  |  | Replication cohort |  |  |
| --- | --- | --- | --- | --- | --- | --- |
| | $\chi^2$<br>Wald $X^2$ | $p$-value | *Odds Ratio (95% CI)* | $\chi^2$<br>Wald $X^2$ | $p$-value | *Odds Ratio (95% CI)* |
|  | *BD/SA*<br>(df = 1) |  |  | *BD/SA*<br>(df = 1) |  |  |
| **Model 8** | 6.302 | **0.012** | 2.36 (1.21-4.62) | 1.663 | **0.020** | 1.15(0.93-1.43) |
| **Model 9** | 5.602 | **0.018** | 2.28 (1.152-4.515) | 3.540 | **0.060** | 1.33(0.99-1.80) |
| **Model 10** | 6.267 | **0.012** | 2.64 (1.235-5.627) | 6.592 | **0.010** | 1.80(1.15-2.83) |
| **Model 11** | 5.692 | **0.017** | 2.30 (1.160-4.565) | 6.311 | **0.010** | 1.12(1.03-1.22) |
| **Model 12** | 5.107 | **0.024** | 2.25 (1.114-4.551) | 5.853 | **0.020** | 1.03(0.66-1.63) |
| **Model 13** | 3.637 | 0.056 | 2.01 (0.981-4.154) | 2.110 | 0.050 | 1.57(1.06-2.33) |

Model 8: dichotomic logistic regression. Model 9: covaried for total number of comorbidities. Model 10: covaried for total number of comorbidities and length of illness. Model 11: covaried for total number of comorbidities, length of illness, and current use of medication. Model 12: covaried for total number of comorbidities, length of illness, current use of medication (any), and current use of lithium. Model 13: covaried for total number of comorbidities, length of illness, current use of medication (any), current use of lithium, and substance use disorder. Nominally significant differences (unadjusted $p < 0.05$ ) are bolded.

## 3. Results

### 3.1 Demographic comparisons

In the discovery cohort (mean age of 36.9 years), BD and CON groups did not significantly differ in chronological age, sex, BMI, and race/ethnicity **(Table 1)**. Their years of education and smoking scores were significantly different, with the BD/SA group having more years of education and higher smoking scores than BD/non-SA and CON (*p*=0.005 and *p*=0.0001, respectively). Age, sex, BMI, race/ethnicity, and years of education were not significantly different between groups in the replication cohort (mean age of 41.9 years); however, BD/SA had a significantly higher smoking score (*p*=0.001). Both cohorts included a similar proportion of male and female subjects, but differed for age, race/ethnicity, smoking scores, and years of education (**Table S2**).

### 3.2 GrimAgeAccel in BD and SA

In our primary analysis, the three groups in the discovery cohort significantly differed on GrimAgeAccel when covaried for age, sex, years of education, and PCs (Model 1: *F*_BD/SA_(2,176)=5.931, *p*=0.003). The group effect remained statistically significant after further adjustment for BMI (Model 2: *F*_BD/SA_(2,164)=5.098, *p*=0.008) and after additional adjustment for white blood cell count proportions (Model 3: *F*_BD/SA_(2,158)=6.227, *p*=0.002) and smoking score (Model 4: *F*_BD/SA_(2,157)=7.904, *p*=0.001). We also found an increased GrimAgeAccel among BD/SA compared to CON when adjusting for smoking score only (Model 5: *F*_BD/SA_(2,188)=6.457, *p*=0.002) (**Table S3**). *Post-hoc* comparisons revealed a significantly greater GrimAgeAccel in BD/SA compared to CON (*p*=0.004, **Figure S2**); based on mean values, this equates to approximately 3 years of accelerated GrimAge. Absolute mean (standard deviation) GrimAgeAccel (in years) was –1.41(4.77) in CON, −0.22(4.87) in BD/non-SA, and +1.42(4.84) in BD/SA (**Table S4)**.

When focusing only on BD groups, GrimAgeAccel was significantly higher in BD/SA compared to BD/non-SA in both the discovery (1.6 years accelerated, *p*=0.046) and replication cohorts (2.2 years accelerated, *p*=0.008) (**Figure 1C-D**; **Tables S4 and 3**). In the discovery cohort, adjusted models showed that BD/SA remained significantly associated with a greater GrimAgeAccel compared to BD/non-SA after adjusting for covariates in Models 1, 3, 4, and 5, but not in Model 2 (*p*=0.051). On the other hand, differences between groups remained significant in all models in the replication cohort, with BD/SA consistently showing higher GrimAgeAccel compared to BD/non-SA (**Table 2**).

### 3.3 Individual GrimAge-associated surrogate and smoking pack-years protein estimates

DNAm-based surrogate protein markers and DNAmPACKYRS significantly correlated with chronological age, with the exception of DNAmLeptin (**Figures S3 and S4)**. An advantage of the GrimAge clock is that each of its surrogate protein DNAm markers can be queried to investigate their role in accelerated aging^25^. Age-adjusted DNAmPAI1 (*p*=0.027) levels and DNAmPACKYRS (*p*=0.031) were significantly higher in BD/SA compared to CON (Model 6, **Table S4**), even after further adjustment for age, sex, PCs, and years of education (*p*=0.003 and *p*=0.017, respectively) (Model 7, **Table S4**). DNAmPAI1 was also elevated in BD/non-SA compared to CON. Additionally, age-adjusted DNAmADM showed a nominally significant difference (*p*=0.039) between BD/SA and CON after covariate adjustment (Model 7, **Table S4**).

When focusing on the two BD subgroups, no significant differences in plasma protein markers were detected in the discovery cohort (**Table 3**). However, in both cohorts, we found a nominally significant difference in BD/SA compared to BD/non-SA on age-adjusted DNAmPACKYRS in Model 6 (discovery *p*=0.046; replication *p*=0.007), which remained after covariate adjustment in Model 7 (*p*=0.034 and *p*=0.007, respectively). In the replication cohort, age-adjusted DNAmLeptin showed a nominally significant difference (*p*=0.024) between BD/SA and BD/non-SA after covariate adjustment (Model 7).

### 3.4 GrimAgeAccel in BD and SA and clinical variables

BD/non-SA and BD/SA significantly differed for many clinical variables in both cohorts (**Table S1**), including MADRS scores, length of illness, total number of comorbidities, and total number of psychiatric hospitalizations. YMRS scores, substance abuse, and age at onset of any mood disorder were also different between groups in the discovery cohort, while current lithium use differed in the replication cohort (**Table S1**). To assess the influence of these clinical variables on the group differences previously identified, we used logistic regression models to test for the association of the diagnostic (BD-non/SA or BD/SA) with the categorical outcome (positive or negative GrimAgeAccel) while adjusting for these clinical variables (**Tables 4 and S5**).

Initially, we found that BD/SA had 69% increased odds of having accelerated GrimAge relative to BD/non-SA, with an odds ratio (OR) of 2.362 (95% CI, 1.207 to 4.623). On the other hand, the BD/non-SA group had 19% lower odds of having accelerated GrimAge relative to BD/SA, with an OR of 0.451 (95% CI, 0.231 to 0.880). These associations remained significant after adjustment for total number of comorbidities (Model 9), comorbidities and length of illness (Model 10), comorbidities, length of illness, and current use of the medication (Model 11), and comorbidities, length of illness, medication, and current lithium use (Model 12). Although the model including substance abuse disorder was not significant (Model 13), the p-value (*p*=0.056) indicates a trend towards an increased odds of accelerated GrimAge for BD/SA compared to BD/non-SA. These associations were replicated in the replication cohort (**Tables 4 and S5)**.

## 4. Discussion

In this study, we extend prior work documenting an accelerated mortality-associated epigenetic clock in BD [22] with novel evidence that SA within BD is associated with even greater GrimAgeAccel. Specifically, we found that BD/SA shows an approximately 3 years higher GrimAgeAccel when compared to CON and at least 1.6 years higher GrimAgeAccel when compared to BD/non-SA. These differences are robust to a variety of potential confounds, including age, sex, years of education, BMI, white blood cell count proportions, genomic ancestry, and smoking. Altogether, these findings pair with existing evidence that BD and SA are associated with accelerated biological aging and provide putative biological mechanisms for premature mortality.

This is the first study to document higher GrimAge in SA compared to a non-psychiatric control group, although previous studies have found accelerated EA related to suicidal behavior [26,50]. SA is typically associated (but not synonymous) with a more severe illness presentation, which may account for some of the observed relationships. SA individuals are more likely to abuse substances [51], report more severe psychiatric symptoms [52] and trauma [53], and come from lower socioeconomic backgrounds [54], which may cumulatively contribute to accelerated aging [16,55-57]. Indeed, BD/SA showed many differences in clinical variables, including longer length of illness and higher numbers of comorbidities and hospitalizations than BD/non-SA. We explored whether the relationship between SA and GrimAgeAccel was attenuated when covarying for these indexes of severity (**Tables 4 and S5**) and found that substance abuse may account for some of the higher GrimAgeAccel observed with SA. Previous studies have documented the association between substance abuse and accelerated aging [58,59].

Medication has also been known to modulate aging, and GrimAge has been positively associated with polypharmacy [20]. When covarying for lithium and medication use separately, the significant difference in GrimAgeAccel between BD groups remained in both cohorts **(Tables 4 and S5)**, suggesting that medication use is not a significant confounder in our results. Nevertheless, we cannot exclude the possibility that other unmeasured factors are responsible for this difference, including participants’ early life or cumulative trauma, which is linked to SA risk [53] and contributes to biological aging [57,60]. SA itself or genetic liability for suicidal behavior may also directly contribute to premature aging, although recent genetic data suggest that EA is largely independent of genetic risk [61].

We further explored the DNAm-based surrogate estimates of seven aging-associated plasma proteins and DNAmPACKYRS used to create the composite GrimAge. In the discovery cohort, analyses revealed that the DNAmPAI-1 was elevated in BD/SA, but even greater in BD/non-SA compared to CON. PAI-1 is a physiological inhibitor of tissue plasminogen activator (tPA) in plasma and is increased in situations related to ischemic cardiovascular events and senescence [62,63]. Recently, increased PAI-1 activity has been linked to mood disorders, with the proposed mechanism involving tPA-mediated cleavage of pro-brain-derived neurotrophic factor (BDNF) into its mature form. Since a lack of tPA or high levels of PAI-1 disrupt BDNF processing, this could result in abnormal neuronal function [64,65] and reduced BDNF levels in BD [66]. Furthermore, plasma levels of BDNF are reduced in suicide attempters [67], and a decreased BDNF expression has also been reported in postmortem brains of suicide decedents [68,69].

In the replication cohort, after covariate adjustment, the DNAmLeptin showed a significant elevation in BD/SA compared to BD/non-SA. Leptin is an adipokine produced in white adipose tissue which crosses the blood-brain barrier [70] to regulate energy homeostasis [71]. An increase in tissue leptin can increase acute inflammatory response [72] and contribute to chronic inflammation in obesity [73]. Indeed, violent SA has been associated with abdominal obesity in BD individuals [74]. However, previous studies have also shown that plasma leptin levels are decreased in suicidal individuals compared to CON [75,76], suggesting that the exact role of this hormone in relation to suicidal behavior warrants further investigation. High levels of leptin are associated with insulin resistance and a higher risk for metabolic syndrome and cardiovascular disease, which have all been previously associated with BD [77,78]. More specifically, older individuals with BD have been shown to present increased arterial stiffness, with evidence of accelerated vascular aging in BD [79]. These findings support the theory of BD as a multisystem illness [80] with several lines of evidence signifying accelerated aging in BD, including shortened telomere length [81] and immunological aging [82].

GrimAgeAccel shows a stepwise elevation from CON to BD/non-SA to BD/SA. Clinically, the most direct implication of this work suggests that interventions targeting factors associated with risk for biological aging may contribute to lowering SA risk or its deleterious biological consequences in BD. Indeed, recent clinical trials suggest that patterns of EA can be reversed with both pharmacological [83] and diet/lifestyle changes [84]. In fact, lithium, which is a known anti-suicidal drug [85], has also been repeatedly shown to present significant anti-aging effects [86]. Moreover, our study also found a difference in the correlation between GrimAge and chronological age, especially for the earliest age group between BD/non-SA and BD/SA in the replication cohort and between BD/SA and controls in the discovery cohort. This suggests that GrimAgeAccel may already be detected in young individuals with a history of SA, which coincides with previous observation of shorter telomere length and lower mitochondrial DNA copy numbers in a subgroup of young BD individuals [87].

Overall, our findings provide robust evidence that the association of SA with GrimAgeAccel in BD is largely resistant to many measures of clinical severity (i.e., this association appears to be specific to SA rather than a more severe clinical presentation of BD). Future studies may consider exploring the association of the GrimAge predictor within specific clinical subtypes in BD, such as suicide decedents in postmortem tissues, and self-injurious thoughts and behaviors. Previous studies have shown associations between accelerated EA and number of SAs [23] as well as SA lethality [26]. Medical comorbidities of BD such as cardiovascular disease [88], asthma [89], diabetes [90], and hypothyroidism [91] may also play a role in EA [92-95]. For now, it is unclear whether psychopathology drives aging or aging drives psychopathology, and there is likely a dynamic interplay between these two processes. Integrating multi-level information in longitudinal studies may be key to uncovering novel understandings of the underlying pathophysiology as well as potential interventional targets.

Limitations of the study include the cross-sectional design and retrospective assessment of SA, which prevents inference about directionality; missing trauma and life stress severity variables; and the small size of the replication sample. Although the replication cohort is more racially homogenous than the multiethnic discovery cohort, we replicated the same between-group findings. Additionally, strengths of our study include the assessment of an independent replication cohort, which validates initial findings and reduces the likelihood of false positives; robust covariates including genomic ancestry, white blood cell count proportions, and an objective DNAm-based smoking score; the inclusion of three groups to tease apart influences of BD and SA; and a rich clinical dataset in both cohorts.

In conclusion, we show that GrimAge, an epigenetic predictor of mortality, shows increasing acceleration from CON to BD/non-SA to BD/SA, and that this acceleration can robustly differentiate BD with and without SA. These relationships are resilient to many possible confounding influences and replicate in an independent cohort, warranting further investigation of EA in suicidal behavior and BD. Considering the severity of both BD symptoms and SA, and the associated reductions in life expectancy, it will be critical to innovate targeted biological tools to reduce morbidity and mortality.

## Data Availability

All data produced in the present study are available upon reasonable request to the authors.

## 5. Acknowledgments

Reagents and sample hybridization in the replication cohort were provided by the Iowa Institute of Human Genetics Genomics Division (IIHG). We would like to thank Hsiang Wen for sample collection and management for the replication cohort. We would like to thank the study participants in both cohorts for their willingness to participate in the study.

## 6. Author Contributions

Design and conceptualization of the study: CNCL, EHCK, MEG, and GRF. Sample collection and recruitment: EHCK, BMRA, JFR, AW, JAW, VAM, JGF, APD, JQ, and JCS. Data generation, processing, and statistical analyses: CNCL, EHCK, SM, ADFC, and APD. Scientific discussion and interpretation of results: CNCL, EHCK, SM, AW, JAW, VAM, APD, JQ, JCS, JGF, MEG, and GRF. Wrote the manuscript: CNCL, EHCK, SM, MEG, and GRF.

