## Supplementary information for "Association between the Epigenetic Lifespan Predictor GrimAge and History of Suicide Attempt in Bipolar Disorder"

**Supplementary figures**

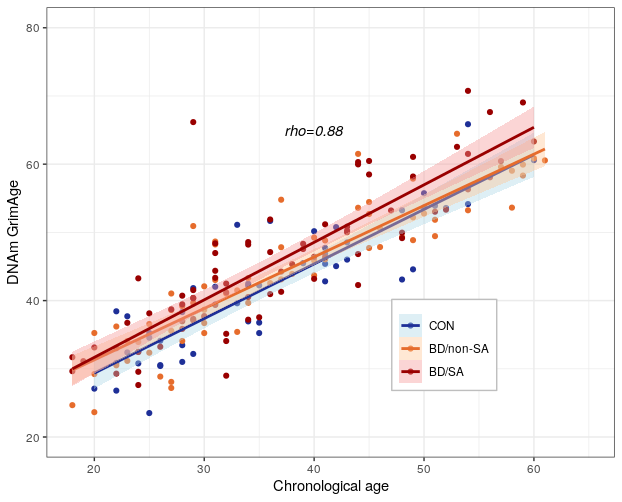

**Figure S1. Correlation between estimated GrimAge and chronological age in the discovery cohort.** Spearman’s correlation analysis indicates a significant correlation (*p<*2.2E-16) between DNA methylation (DNAm) GrimAge and chronological age in the entire sample, as well as in the subgroups (CON: rho=0.89, *p*<2.2E-16; BD/non-SA: rho=0.91, *p*<2.2E-16; BD/SA: rho=0.85, *p*<2.2E-16).

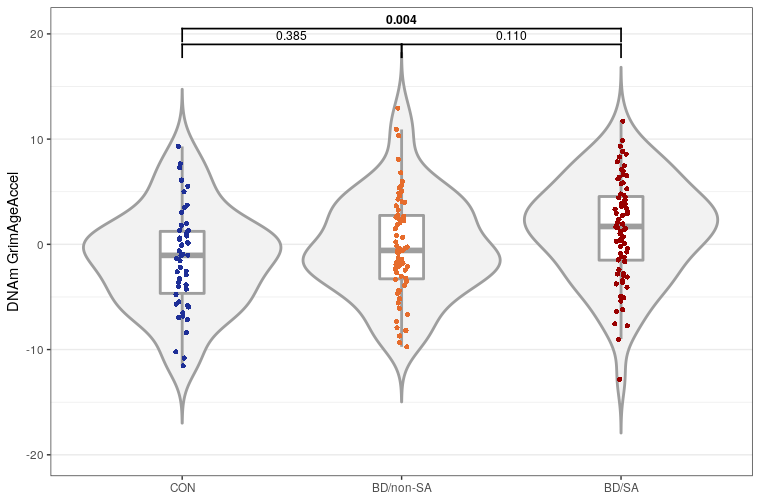

**Figure S2. Violin plots with dots showing measures of DNA methylation (DNAm) accelerated GrimAge (GrimAgeAccel, in years) in the discovery cohort.** ANOVA with Tukey's *post hoc* test was performed for comparisons between the BD/SA (n=67), BD/non-SA (n=77), and CON (n=50). BD/SA, bipolar disorder with history of suicide attempt. BD/non-SA, bipolar disorder with no history of suicide attempt. CON, non-psychiatric control.

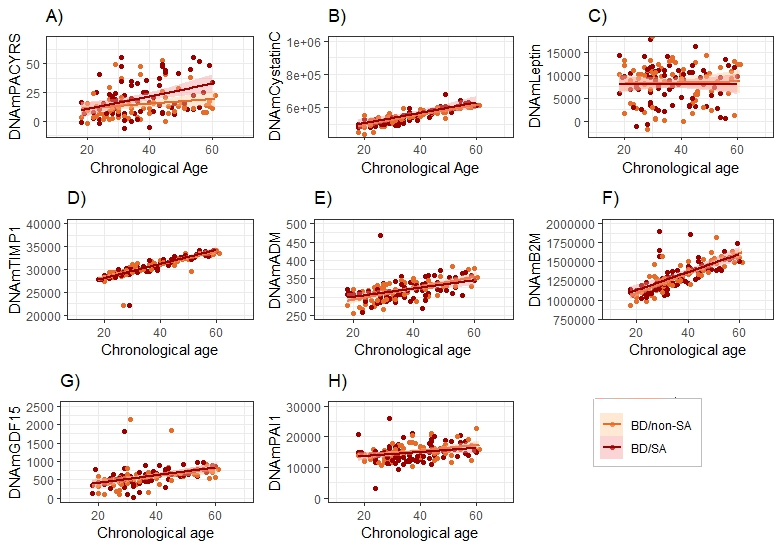

**Figure S3. Chronological age versus GrimAge clock elements in the discovery cohort.** Chronological age versus A) PACKYRS (rho=0.31, *p*=1.9E-04); BD/non-SA (rho=0.30, *p*=0.014); BD/SA (rho=0.34, *p*=0.002); B) Cystatin C (rho=0.86, *p*<2.2E-16); BD/non-SA (rho=0.87, *p*<2.2E-16); BD/SA (rho=0.84, *p*<2.2E-16); C) Leptin (rho=0.03, *p*=0.753); BD/non-SA (rho=0.04, *p*=0.789); BD/SA (rho=0.04, *p*=0.761); D) TIMP1 (rho=0.93, *p*<2.2E-16); BD/non-SA (rho=0.93, *p*<2.2E-16); BD/SA (rho=0.94, *p*<2.2E-16); E) ADM (rho=0.57, *p*<6.62E-14); BD/non-SA (rho=0.65, *p*=2.6E-09); BD/SA (rho=0.50, *p*=3.3E-06); F) B2M (rho=0.79, *p*<2.2E-16); BD/non-SA (rho=0.84, *p*<2.2E-16); BD/SA (rho=0.76, *p*=1.4E-15); G) GDF-15 (rho=0.62, *p*<2.2E-16); BD/non-SA (rho=0.69, *p*=1.2E-10); BD/SA (rho=0.59, *p*=1.9E-08); and H) PAI-1 (rho=0.33, *p*=5.0E-05); BD/non-SA (rho=0.45, *p*= 1.6E-04); BD/SA (rho=0.24, *p*=0.035).

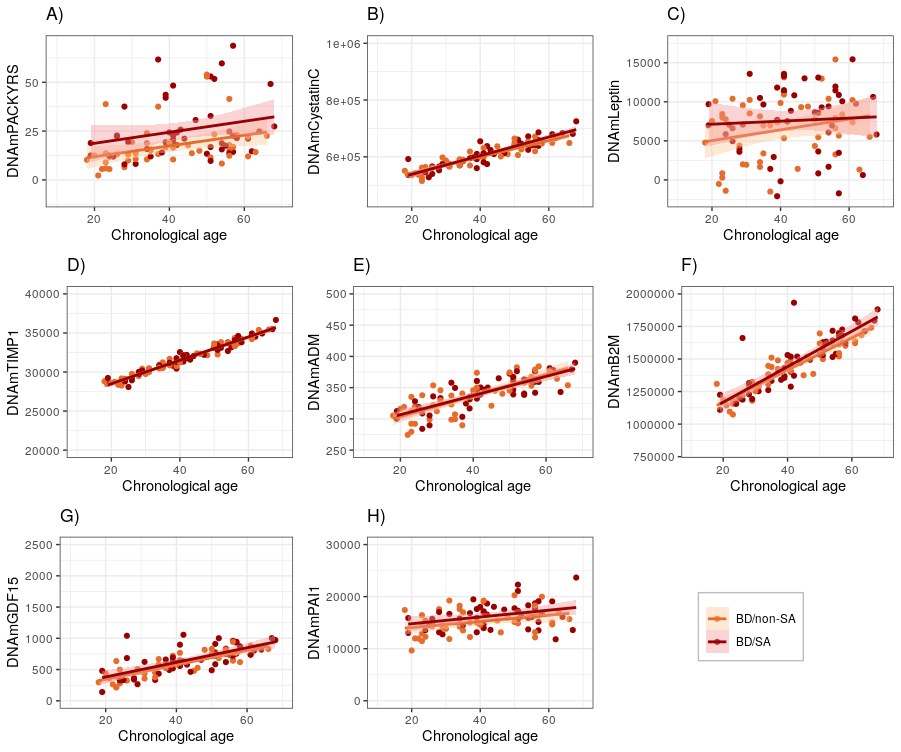

**Figure S4. Chronological age versus GrimAge clock elements in the replication cohort.** Chronological age versus A) PACKYRS (rho=0.41, *p*=4.67E-05); BD/non-SA (rho=0.49, *p*=4.5E-04); BD/SA (rho=0.28, *p*=0.058); B) Cystatin C (rho=0.91, *p*<2.2E-16); BD/non-SA (rho=0.94, *p*<2.2E-16); BD/SA (rho=0.91, *p*<2.2E-16) C) Leptin (rho=0.18, *p*=0.088); BD/non-SA (rho=0.18, *p*=0.214); BD/SA (rho=0.11, *p*=0.465); D) TIMP1 (rho=0.98, *p*<2.2E-16); BD/non-SA (rho=0.98, *p*<2.2E-16); BD/SA (rho=0.97, *p*<2.2E-16); E) ADM (rho=0.81, *p*<2.2E-16); BD/non-SA (rho=0.80, *p*=1.2E-11); BD/SA (rho=0.79, *p*=2.9E-11); F) B2M (rho=0.89, *p*<2.2E-16); BD/non-SA (rho=0.92, *p*<2.2E-16); BD/SA (rho=0.86, *p*=4.8E-15); G) GDF-15 (rho=0.77, *p*<2.2E-16); BD/non-SA (rho=0.85, *p*=6.2E-14); BD/SA (rho=0.69, *p*=7.3E-08); and H) PAI-1 (rho=0.33, *p*=0.001); BD/non-SA (rho=0.36, *p*=0.014); BD/SA (rho=0.31, *p*=0.033).

**Table S1. Clinical characteristics between BD/non-SA and BD/SA groups.**

|  | **Discovery cohort** | | | **Replication cohort** | | |
| --- | --- | --- | --- | --- | --- | --- |
|  | **BD/non-SA (*n*=67)** | **BD/SA (*n*=77)** | ***p*-value** | **BD/non-SA (*n*=47)** | **BD/SA (*n*=48)** | ***p*-value** |
| YMRS total **^3^**  mean (SD) | 5.20 (7.00) | 7.10 (8.08) | **0.022** | 5.36 (5.46) | 8.21 (8.31) | 0.070 |
| MADRS total **^3^**  mean (SD) | 10.47 (1.21) | 16.35 (1.15) | **3.03E-04** | 11.9(8.65) | 16.2 (8.95) | **0.022** |
| Length of illness**^3^**  mean (SD) | 11.6 (10.3) | 15.0 (10.6) | **0.038** | 18.6 (13.0) | 23.8 (13.1) | **0.045** |
| Comorbidities total**^3^**  mean (SD) | 1.55 (1.64) | 2.16 (1.62) | **0.007** | 1.43 (1.81) | 2.46 (1.87) | **0.002** |
| Substance abuse **^1^**  n (%) | 32 (47.8%) | 53 (68.8%) | **0.010** | 2 (4.3%) | 6 (12.5%) | 0.268 |
| Total hospitalization**^2^**  mean (SD) | 2.46 (2.64) | 4.75 (5.50) | **0.005** | 2.72 (3.84) | 7.02 (7.49) | **1.01E-04** |
| Age onset of any mood disorder **^2^**  mean (SD) | 21.9 (23.4) | 21.6 (7.98) | **0.026** | 21.7 (8.65) | 20.7 (10.8) | 0.175 |
| Current use of lithium **^1^**   n (%) | 16 (23.9%) | 15 (19.5%) | 0.548 | 20 (42.6%) | 6 (12.5%) | **0.001** |
| Current use of medication **^1^**   n (%) | 59 (88.1%) | 69 (75.3%) | 0.796 | 42 (89.4%) | 44 (91.7%) | 0.740 |

^1^Fisher's exact test. ^2^t-test. ^3^Mann-Whitney/Wilcoxon rank sum test with continuity correction. BD/SA, bipolar disorder with history of suicide attempt. BD/non-SA, bipolar disorder with no history of suicide attempt. Nominally significant differences (*p*<0.05) are bolded. MADRS - Montgomery-Åsberg Depression Rating Scale; YMRS – Young Mania Rating Scale.

**Table S2. Demographic and clinical characteristics of bipolar disorder groups in the discovery and replication cohorts**

|  | **Discovery Cohort** | **Replication Cohort** | ***p*-value** |
| --- | --- | --- | --- |
| **Demographic and clinical characteristics** | | | |
| Female^1^  n (%) | 103 (71.5%) | 62 (65.3%) | 0.320 |
| Age^2^  mean (SD) | 36.9 (11.3) | 41.9 (13.5) | **0.005** |
| Race^1^  *White*  *Hispanic/Latinx*  *Black/African American*  *American Indian/Pacific Islander*  *Asian*  *More than one race*  Missing | 58 (40.3%)  22 (15.3%)  47 (32.6%)  2 (1.39%)  0 (0.0%)  10 (6.9%)  5 | 74 (77.9%)  9 (9.5%)  4 (4.2%)  2 (2.1%)  1 (1.1%)  3 (3.2%)  2 | **1.65E-09** |
| Smoking score^2^  mean (SD) | 4.00 (1.32) | 3.24 (4.35) | **1.21E-07** |
| Years of education^2^  mean (SD) | 14.0 (2.50) | 15.0 (2.21) | **0.005** |
| BMI^2^  mean (SD) | 30.1 (7.49) | 30.7 (7.40) | 0.532 |
| **White blood cells** |  |  |  |
| CD8T^2^  mean (SD) | 0.010 (0.061) | 0.088 (0.045) | 0.091 |
| CD4T^2^  mean (SD) | 0.130 (0.077) | 0.112 (0.049) | **0.028** |
| NK^2^  mean (SD) | 0.021 (0.034) | 0.006 (0.017) | **0.004** |
| Mono  mean (SD) | 0.056 (0.033) | 0.063 (0.025) | 0.065 |
| B cell^2^  mean (SD) | 0.031 (0.037) | 0.010 (0.021) | **1.01E-12** |
| Gran^2^  mean (SD) | 0.582 (0.175) | 0.567 (0.086) | 0.178 |
| **Clinical variables** |  |  |  |
| YMRS total^2^  mean (SD) | 6.24 (7.62) | 6.80 (7.15) | **1.63E-15** |
| MADRS total^2^  mean (SD) | 13.6 (10.4) | 14.0 (9.02) | **2.20E-16** |
| Length of illness^2^  mean (SD) | 13.4 (10.6) | 21.2 (13.3) | **2.20E-16** |
| Comorbidities total^2^  mean (SD) | 1.87 (1.65) | 1.95 (1.90) | 0.864 |
| Substance abuse^1^  n (%) | 85 (59.0%) | 8 (8.4%) | **2.20E-16** |
| Total hospitalization^2^  mean (SD) | 3.69 (4.54) | 4.89 (6.32) | **1.31E-08** |
| Age onset of any mood disorder^2^  mean (SD) | 23.5 (8.96) | 21.2 (9.74) | **2.20E-16** |
| Current use of lithium^1^  mean (SD) | 31 (21.5%) | 26 (27.4%) | 0.353 |
| Current use of medication^1^  n (%) | 128 (88.9%) | 86 (90.5%) | 0.830 |

^1^Fisher’s Exact Test for Count Data. ^2^Welch Two Sample t-test/Wilcoxon rank sum test with continuity correction. Nominally significant differences (unadjusted *p*<0.05) are bolded. B cell – B lymphocytes; CD4+ T-lymphocytes; CD8T – CD8+ T-lymphocytes; Gran – granulocytes; MADRS - Montgomery-Åsberg Depression Rating Scale; Mono – monocytes; NK – natural killer cells; SD – standard deviation; YMRS – Young Mania Rating Scale.

**Table S3. Adjusted analysis of differences in GrimAgeAccel between CON, BD/non-SA, and BD/SA groups.**

|  | **Discovery cohort** | |
| --- | --- | --- |
|  | ***F*_BD/SA_ (df,df = 2, )** | ***p-*value** |
| **Model 1** | 5.931 | **0.003** |
| **Model 2** | 5.098 | **0.008** |
| **Model 3** | 6.227 | **0.002** |
| **Model 4** | 7.904 | **0.001** |
| **Model 5** | 6.457 | **0.002** |

Model 1: covaried for age, sex, GWAS principal components (PCs), and years of education. Model 2: covaried for age, sex, GWAS PCs, years of education, and body mass index (BMI). Model 3: covaried for sex, GWAS PCs, years of education, BMI, and white blood cell proportions. Model 4: covaried for age, sex, GWAS PCs, years of education, BMI, white blood cell proportions, and smoking score. Model 5: covaried for smoking score. Bonferroni-corrected *p*<0.01 was taken as the significance threshold. Nominally significant differences (unadjusted *p*<0.05) are bolded. BD/SA, bipolar disorder with history of suicide attempt. BD/non-SA, bipolar disorder with no history of suicide attempt. CON, non-psychiatric controls.

**Table S4. GrimAge clock and its components between CON, BD/non-SA, and BD/SA groups in the discovery cohort.**

|  | **Discovery cohort** | | | | | | |
| --- | --- | --- | --- | --- | --- | --- | --- |
|  | **CON**  **(*n* =50)** | **BD/non-SA (n= 67)** | **BD/SA**  **(n =77)** | **Model 6** | | **Model 7** | |
|  | **Mean**  **(SD)** | **Mean**  **(SD)** | **Mean**  **(SD)** | ***F*_BD/SA_ (df, df = 2,)** | ***p*-value** | ***F*_BD/SA_ (df, df = 2,)** | ***p*-value** |
| **GrimAgeAccel** | -1.41 (4.77) | -0.217 (4.87) | 1.42 (4.84) | 5.424 | **0.005** | 5.931 | **0.003** |
| **DNAmPACKYRS** | -2.34 (10.9) | -1.25 (12.5) | 3.18 (13.7) | 3.543 | **0.031** | 4.185 | **0.017** |
| **DNAmCystatinC** | 5940 (57098) | -6331 (43894) | 532 (40552) | 1.011 | 0.366 | 1.012 | 0.366 |
| **DNAmLeptin** | -85.0 (4397) | 67.7 (4246) | -166 (3895) | 0.057 | 0.945 | 0.207 | 0.814 |
| **DNAmTIMP1** | -165 (1061) | 76.3 (875) | 101 (854) | 1.415 | 0.246 | 1.399 | 0.250 |
| **DNAmADM** | -5.37 (24.8) | 2.28 (22.8) | 0.944 (23.3) | 1.642 | 0.196 | 3.306 | **0.039** |
| **DNAmB2M** | -14708 (158638) | 5609 (134347) | 7723 (145708) | 0.396 | 0.674 | 0.506 | 0.604 |
| **DNAmGDF15** | -55.1 (244) | -23.3 (244) | 43.4 (271) | 2.472 | 0.087 | 2.699 | 0.072 |
| **DNAmPAI1** | -710 (2185) | 579 (2460) | 17.9 **(**2755) | 3.685 | **0.027** | 3.856 | **0.023** |

Model 6: ANOVA. Model 7: covaried for age, sex, GWAS principal components (PCs), and years of education. Bonferroni-corrected *p*<0.005 was taken as the significance threshold. Nominally significant differences (*p*<0.05) are bolded. BD/SA, bipolar disorder with history of suicide attempt. BD/non-SA, bipolar disorder with no history of suicide attempt. CON, non-psychiatric controls.

**Table S5. Phenotype-adjusted differences in GrimAgeAccel between BD/non-SA and BD/SA groups.**

|  | **Discovery cohort** | | | **Replication cohort** | | |
| --- | --- | --- | --- | --- | --- | --- |
|  | **_Wald_ *_X_^2^ _BD/non-SA_***  **(df = 1)** | ***p*-value** | ***Odds Ratio (95% CI)*** | **_Wald_ *_X_^2^ _BD/non-SA_***  **(df = 1)** | ***p*-value** | ***Odds Ratio (95% CI)*** |
| **Model 8** | 5.452 | **0.020** | 0.451 (0.231-0.880) | 9.567 | **0.001** | 0.08(0.74-0.90) |
| **Model 9** | 5.602 | **0.018** | 0.438 (0.222-0.868) | 6.18 | **0.010** | 0.73(0.56-0.95) |
| **Model 10** | 4.795 | **0.029** | 0.459 (0.228-0.922) | 8.19 | **0.002** | 0.64(0.48-0.86) |
| **Model 11** | 5.692 | **0.017** | 0.434 (0.219-0.862) | 3.62 | **0.030** | 0.89(0.71-1.35) |
| **Model 12** | 5.107 | **0.024** | 0.444 (0.220-0.898) | 3.54 | **0.045** | 1.10(0.87-1.39) |
| **Model 13** | 3.637 | 0.056 | 0.495 (1.165-6.507) | 3.03 | 0.080 | 0.96(0.92-1.01) |

Model 8: dichotomic logistic regression. Model 9: covaried for total number of comorbidities. Model 10: covaried for total number of comorbidities and length of illness. Model 11: covaried for total number of comorbidities, length of illness, and current use of medication (any). Model 12: covaried for total number of comorbidities, length of illness, current use of medication (any), and current use of lithium. Model 13: covaried for total number of comorbidities, length of illness, current use of medication (any), current use of lithium, and substance use disorder. Nominally significant differences (unadjusted *p*<0.05) are bolded.
